# Predicting post-stroke cognitive impairment using electronic health record data

**DOI:** 10.1101/2024.02.02.24302240

**Authors:** Jeffrey M. Ashburner, Yuchiao Chang, Bianca Porneala, Sanjula D. Singh, Nirupama Yechoor, Jonathan M. Rosand, Daniel E. Singer, Christopher D. Anderson, Steven J. Atlas

## Abstract

**Importance:** Secondary prevention interventions to reduce post-stroke cognitive impairment (PSCI) can be aided by the early identification of high-risk individuals who would benefit from risk factor modification.

**Objective:** To develop and evaluate a predictive model to identify patients at increased risk of PSCI over 5 years using data easily accessible from electronic health records.

**Design:** Cohort study with patients enrolled between 2003-2016 with follow-up through 2022.

**Setting:** Primary care practices affiliated with two academic medical centers.

**Participants:** Individuals 45 years or older, without prior stroke or prevalent cognitive impairment, with primary care visits and an incident ischemic stroke between 2003-2016 (development/internal validation cohort) or 2010-2022 (external validation cohort).

**Exposures:** Predictors of PSCI were ascertained from the electronic health record.

**Main Outcome:** The outcome was incident dementia/cognitive impairment within 5 years and beginning 3 months following stroke, ascertained using ICD-9/10 codes. For model variable selection, we considered potential predictors of PSCI and constructed 400 bootstrap samples with two-thirds of the model derivation sample. We ran 10-fold cross-validated Cox proportional hazards models using a least absolute shrinkage and selection operator (LASSO) penalty. Variables selected in >25% of samples were included.

**Results:** The analysis included 332 incident diagnoses of PSCI in the development cohort (n=3,741), and 161 and 128 incident diagnoses in the internal (n=1,925) and external (n=2,237) validation cohorts. The c-statistic for predicting PSCI was 0.731 (95% CI: 0.694-0.768) in the internal validation cohort, and 0.724 (95% CI: 0.681-0.766) in the external validation cohort. A risk score based on the beta coefficients of predictors from the development cohort stratified patients into low (0-7 points), intermediate (8-11 points), and high (12-35 points) risk groups. The hazard ratios for incident PSCI were significantly different by risk categories in internal (High, HR: 6.2, 95% CI 4.1-9.3; Intermediate, HR 2.7, 95% CI: 1.8-4.1) and external (High, HR: 6.1, 95% CI: 3.9-9.6; Intermediate, HR 2.8, 95% CI: 1.9-4.3) validation cohorts.

**Conclusions and Relevance:** Five-year risk of PSCI can be accurately predicted using routinely collected data. Model output can be used to risk stratify and identify individuals at increased risk for PSCI for preventive efforts.

## INTRODUCTION

The burden of stroke-related disability and death on patients, their families and the general public is immense.^1^ Stroke survivors are at increased risk of acquired cognitive impairment and dementia compared to those without stroke.^2^ Post-stroke cognitive impairment (PSCI) may range from mild to severe and may impact up to 60% of stroke survivors.^3^ Though the highest risk of PSCI occurs shortly after the stroke event, many develop new or progressive cognitive impairment or dementia several months to years after the stroke event.^3^ Unmodified risk factors account for a substantial proportion of the risk for PSCI.^4^ An efficient, automated alerting system that identifies those who would benefit from risk factor modification could have substantial public health impact.^5–7^ Secondary prevention interventions to reduce the incidence of PSCI can be aided by the early identification of high-risk individuals and a focus on brain health and brain care.^8^

Existing models to predict PSCI have demonstrated moderate accuracy but are not amenable to large-scale prospective implementation. Published models have largely focused on short-term follow-up (< 6 months) following stroke,^9–11^ and often include predictors that are not available or easily accessible in electronic health records (EHRs).^10–14^ Models that predict PSCI over longer periods may be more useful clinically to allow time for secondary prevention efforts.

Additionally, prediction models that can be implemented and fully automated within the EHR can assess risk at a population level and help facilitate efficient identification and enrollment of high-risk patients into preventive interventions.

In this study, we developed and evaluated a predictive model to identify post-stroke patients at increased risk of PSCI over 5 years using data easily accessible in EHRs. Our population included patients within two academic primary care networks without a prior history of stroke or cognitive impairment at cohort enrollment.

## METHODS

### Study Sample

#### Model Development and Internal Validation

Study cohorts for model development and internal validation consisted of patients from the Primary Care Practice-Based Research Network at Massachusetts General Hospital (MGH) identified using a validated attribution algorithm.^15,16^ All 18 practices in the network use a common EHR and data warehouse for records from 7 affiliated Mass General Brigham (MGB) hospitals including MGH and Brigham and Women’s Hospital (BWH).^17^ Individuals included were 45 years or older who had primary care visits and experienced an incident ischemic stroke event between 2003-2016. Patients excluded were those with prevalent cognitive impairment or prior history of ischemic or hemorrhagic stroke at the time of cohort enrollment. Stroke events were defined based on two ICD-9/10 codes for stroke on different dates, with the earliest date considered as the stroke date (**Table S1**). The cohort was randomly split into two subsets: two-thirds for model development and one-third for internal validation.

#### External Validation

The external validation cohort consisted of patients 45 years or older from 15 BWH primary care practices with visits and experienced an incident ischemic stroke event between 2010-2022, and did not have prevalent cognitive impairment or a history of stroke prior to cohort enrollment. This medical records-based study was approved with a waiver of informed consent by the local MGB Institutional Review Board. MGB data contains protected health information and cannot be shared publicly. The data processing scripts used to perform analyses will be made available to interested researchers upon reasonable request to the corresponding author.

### Ascertainment of Potential Predictors from EHR

Data extracted from the EHR included 1) patient demographics, 2) diagnostic codes (ICD-9 and ICD-10), 2) procedure codes (Current Procedural Terminology [CPT]), 3) medications, 4) laboratory values, 5) unstructured progress/visit notes, 6) unstructured reports, and 7) vital status. We assembled a list of potential predictors of PSCI based on a review of prior studies. The full list of potential model features is available in **Table S2**.

### Clinical Characteristics

Patient characteristics and comorbidities were ascertained using EHR data from prior to the date of cohort entry for each patient, which was 3 months after the stroke date. Age, sex, race/ethnicity, education, marital status, and insurance status were ascertained at the time of cohort entry. Height and weight recorded closest to cohort entry were obtained for body mass index calculations. Medication use was assessed based on any medications listed in the EHR in the 3 years prior to cohort entry. Polypharmacy was defined as 5 different medications listed in the EHR in the prior 3 years. Laboratory values were assessed within 3 years prior to cohort entry and grouped into standard categories. Patients without laboratory values were assumed to be normal. Current smoking status was assessed using the most recent smoking status update in the EHR prior to cohort entry. We used patient home zip code to link to the National Neighborhood Data Archive and ascertain the percentage of the population within a zip code tabulation area with household income less than the poverty level, $30,000, and $50,000.^18^ We considered individual components of an electronic frailty index (eFI) as potential predictors (n=35 of 36 predictors considered, excluding ‘memory problems’).^19^ Sedentary physical activity level was ascertained by searching progress and visit notes for key words and phrases, including: “sedentary,” “discussed weight loss and exercise,” “discussed benefits of exercise,” “encouraged physical activity,” and “encouraged exercise.” Discharge disposition from an MGB hospital to a facility (assisted living, nursing home, acute rehabilitation) was ascertained from discharge summaries. Evidence of ventricular enlargement, white matter disease, and white matter infarcts were defined based on regular expression coding of brain MRI reports anytime prior to cohort entry (**Table S3**).

We utilized validated EHR algorithms to define the following variables: obesity, diabetes mellitus, hypertension, congestive heart failure, coronary artery disease, peripheral vascular disease, cerebrovascular disease, atrial fibrillation, chronic kidney disease.^20–24^ Comorbidities without a validated algorithm were identified by a single ICD-9/10 code prior to and within 3 years of cohort entry. Sleep disturbances were defined by a single ICD-9/10 code or multiple refills of benzodiazepines marketed for sleep (estazolam, Prosom, flurazepam, Dalmane, temazepam, Restoril, quazepam, Doral, triazolam, Halcion) within 3 years prior to cohort entry.

### Outcomes

The primary outcome was incident dementia/cognitive impairment within 5 years following a first stroke event. Follow-up began 3 months after the stroke event to exclude diagnoses of cognitive impairment directly associated with the stroke. Incidence of PSCI was based on two ICD-9/10 codes for dementia or cognitive impairment following cohort entry (3 months after date of stroke event) (**Table S4**).^25^ Cases included in analyses occurred between 2003-2021 for the development and internal validation cohorts, and between 2010-2022 for the external validation cohort.

### Model Derivation

In the development cohort, we used Cox proportional hazards models to predict PSCI within 5 years (with follow-up beginning 3 months following incident stroke). Censoring occurred at time of death, last primary care visit if leaving the primary care cohort, or after the end of the follow-up period (5 years). For variable selection we considered all potential model features in **Table S2** and constructed 400 bootstrap samples with two-thirds of the model derivation sample. We ran 10-fold cross-validated Cox proportional hazards models using a least absolute shrinkage and selection operator (LASSO) penalty.^26^ The largest tuning parameter (lambda) was selected such that error was within 1 standard deviation of the minimum cross-validated error.^27^ Variables that were selected in >25% of samples were included in the final model. Within each model, individual 5-year risk of the outcome was calculated.

### Statistical Analysis

In validation cohorts, we used the coefficients from the model established in the development cohort to estimate predicted 5-year risks and used the product-limit method, which accounts for censoring, to calculate observed 5-year risks. In the external validation cohort, administrative censoring occurred at the end of follow-up in 2022. We calculated Harrell’s C-statistic to assess the model’s ability to separate who developed incident cognitive impairment from those who did not. Variables from the final multivariable Cox regression model were converted to a risk score, with points assigned to each predictor approximately proportional to the magnitude of the regression coefficients rounded to the nearest integer. The risk score was categorized into “low,” “intermediate,” and “high” risk groups based on the observed rate of incident cognitive impairment. We chose thresholds in our point score that appeared to optimally aggregate low- and high-risk groups. We evaluated event rates, hazard ratios (HR), and cumulative incidence curves stratified by risk groups. We considered a two-sided p value <0.05 to indicate statistical significance.

## RESULTS

The study sample included 5,666 patients aged 45 years or older who had a first ischemic stroke and did not have prevalent cognitive impairment. This sample was randomly split into a development cohort (n=3,741) and internal validation cohort (n=1,925). The mean age in the development cohort was 71.4 years (SD 11.8 years), 50.5% were female, and 79.3% were non-Hispanic White. The mean age in the internal validation cohort was 71.2 years (SD 11.8 years), 49.0% were female, and 81.2% were non-Hispanic White. The external validation cohort included 2,237 patients meeting eligibility criteria (mean age 67.1 years, 50.5% female, 66.2% non-Hispanic White) (**Table 1**).

**Table 1:** Baseline patient characteristics for development, internal validation, and external validation cohorts.

|  | Development<br>(n=3,741) | Internal Validation<br>(n=1,925) | External Validation<br>(n=2,237) |
| --- | --- | --- | --- |
| Age, mean (SD) | 71.4 (11.8) | 71.2 (11.8) | 67.0 (11.3) |
| 45-55 years | 356 (9.5%) | 208 (10.8%) | 356 (15.9%) |
| 55-65 years | 794 (21.2%) | 351 (18.2%) | 566 (25.3%) |
| 65-75 years | 1051 (28.1%) | 571 (29.7%) | 711 (31.8%) |
| 75-85 years | 1034 (27.6%) | 559 (29.0%) | 463 (20.7%) |
| ≥ 85 years | 506 (13.5%) | 236 (12.3%) | 141 (6.3%) |
| Sex, female | 1872 (50.5%) | 944 (49.0%) | 1126 (50.3%) |
| Race/ethnicity |  |  |  |
| Non-Hispanic White | 2965 (79.3%) | 1563 (81.2%) | 1472 (65.8%) |
| Black | 256 (6.8%) | 103 (5.4%) | 360 (16.1%) |
| Hispanic | 70 (1.9%) | 40 (2.1%) | 165 (7.4%) |
| Asian | 187 (5.0%) | 92 (4.8%) | 44 (2.0%) |
| Other | 141 (3.8%) | 86 (4.5%) | 130 (5.8%) |
| Unknown | 122 (3.3%) | 41 (2.1%) | 66 (3.0%) |
| Insurance |  |  |  |
| Commercial | 1190 (31.8%) | 589 (30.6%) | 938 (41.9%) |
| Medicare | 2036 (54.4%) | 1076 (55.9%) | 1189 (53.2%) |
| Medicaid | 391 (10.5%) | 201 (10.4%) | 109 (4.9%) |
| Self-pay | 124 (3.3%) | 59 (3.1%) | 1 (0.0%) |
| % with Income < poverty level by zip code, mean (SD)* | 11.5% (7.6%) | 11.1% (7.5%) | 11.8% (9.7%) |
| Mobility problems | 1385 (37.0%) | 706 (36.7%) | 850 (38.0%) |
| Prior falls | 1307 (34.9%) | 677 (35.2%) | 493 (22.0%) |
| Delirium | 751 (20.1%) | 396 (20.6%) | 265 (11.9%) |
| Peripheral vascular disease | 521 (13.9%) | 234 (12.2%) | 135 (6.0%) |
| Parkinson's disease | 379 (10.1%) | 196 (10.2%) | 191 (8.5%) |
| Depression | 623 (16.7%) | 318 (16.5%) | 718 (32.1%) |
| Chronic kidney disease, severe | 306 (8.2%) | 149 (7.7%) | 140 (6.3%) |
| Abnormal weight loss and anorexia | 928 (24.8%) | 449 (23.3%) | 398 (17.8%) |
| Discharge to facility | 1133 (30.3%) | 553 (28.7%) | 365 (16.3%) |

\* Ascertained from National Neighborhood Data Archive (NaNDA).<sup>18</sup>

### Development Cohort

In the development cohort, there were 332 incident diagnoses of PSCI (5-year Kaplan-Meier cumulative incidence, 11.4%; 95% CI: 10.3-12.7%) and 241 death events that occurred prior to a diagnosis of cognitive impairment or the end of 5 years of follow-up (12.5%). Among those who developed PSCI, the median time from the date of incident stroke to a diagnosis of PSCI was 2.0 years (IQR 0.9 – 3.3 years). Variables selected in the final model included age, insurance, mobility problems, prior history of falls, delirium, peripheral vascular disease, Parkinson’s disease, depression, severe chronic kidney disease, abnormal weight loss and anorexia, and discharge from the hospital to a facility. The estimated beta coefficients and hazard ratios for variables included in the final model are shown in **Table 2**. Based on the final model, we assigned points proportional to the beta coefficients to create a risk scheme with a possible range of 0 to 35 points (**Table 2**). We considered those with 0-7 points as “low” risk, 8-11 points as “medium” risk, and 12-35 points as “high” risk. The C-statistic using the risk score to predict PSCI was 0.750 (95% CI: 0.726-0.775). The observed rates of incident PSCI by risk groups are shown in **Table 3**. HRs for incident cognitive impairment by low, intermediate, and high risk groups are shown in **Table 4**. Cumulative incidence plots stratified by risk groups are shown in **Figure 1**.

**Figure 1:**
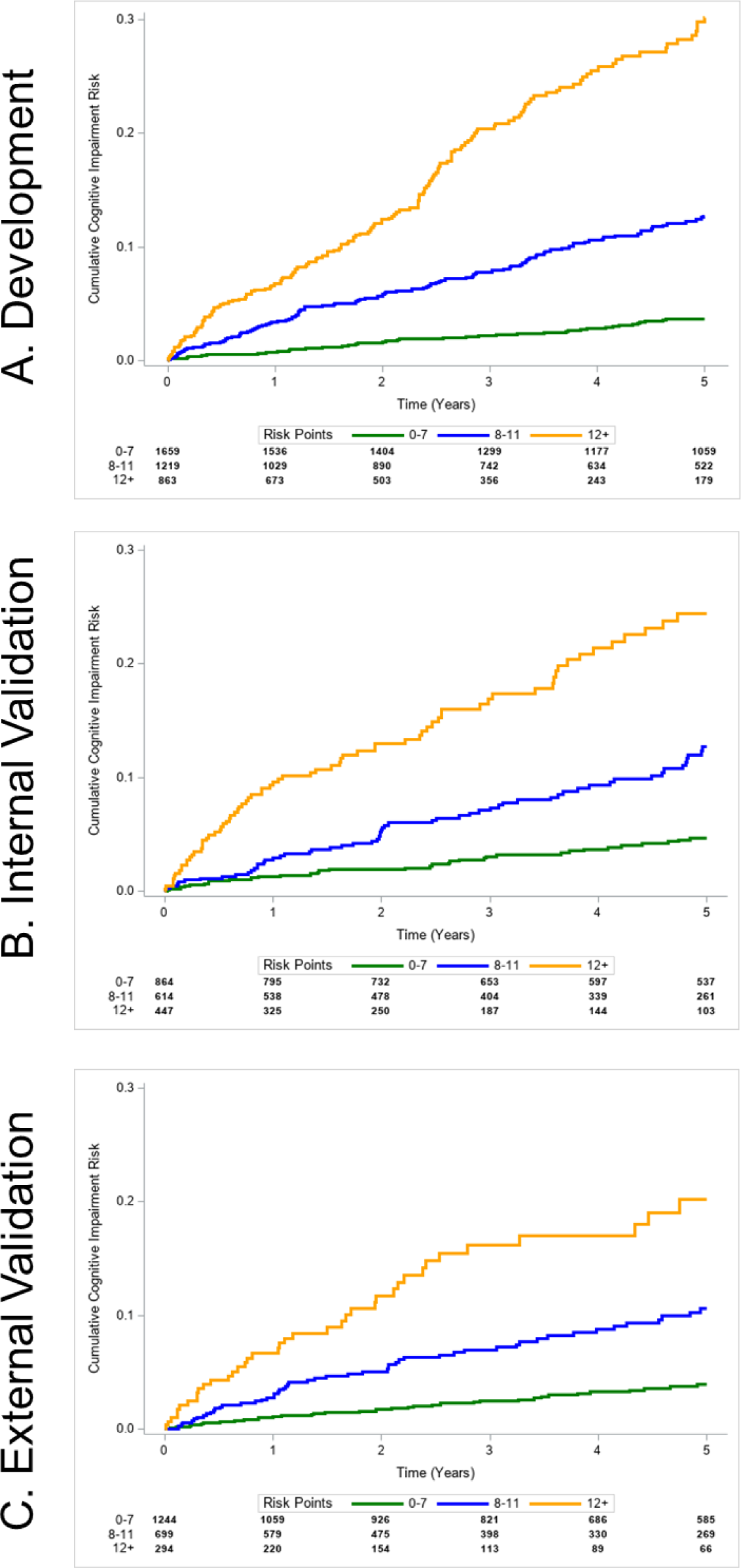
Cumulative incidence plots stratified by low-, intermediate-, and high-risk groups in the development (Panel A), internal validation (Panel B), and external (Panel C) validation cohorts.

**Table 2:** Estimated beta coefficients and hazard ratios for features included in the post-stroke cognitive impairment prediction model in the development cohort*.

| | Estimated $\beta$ (SE) | HR (95% CI) | Points |
| --- | --- | --- | --- |
| Age, yr |  |  |  |
| 45 - < 55 | - | - | 0 |
| 55 - < 65 | 0.108 (0.371) | 1.11 (0.54-2.31) | 1 |
| 65 - < 75 | 0.791 (0.347) | 2.21 (1.12-4.35) | 4 |
| 75 - < 85 | 1.310 (0.349) | 3.71 (1.87-7.34) | 6 |
| $\geq 85$ | 1.715 (0.360) | 5.56 (2.75-11.25) | 8 |
| Insurance |  |  |  |
| Medicaid | 0.236 (0.268) | 1.27 (0.75-2.14) | 1 |
| Medicare | 0.420 (0.162) | 1.52 (1.11-2.09) | 2 |
| Non-Government | - | - | 0 |
| Mobility problems | 0.410 (0.121) | 1.51 (1.19-1.91) | 2 |
| Prior falls | 0.312 (0.118) | 1.37 (1.09-1.72) | 1 |
| Delirium | 0.347 (0.130) | 1.42 (1.10-1.82) | 2 |
| Peripheral vascular disease | 0.339 (0.133) | 1.40 (1.08-1.82) | 2 |
| Parkinson's disease | 0.227 (0.156) | 1.26 (0.93-1.70) | 1 |
| Depression | 0.490 (0.135) | 1.63 (1.25-2.13) | 2 |
| Chronic kidney disease, severe | 0.205 (0.176) | 1.23 (0.87-1.73) | 1 |
| Abnormal weight loss and anorexia | 0.189 (0.119) | 1.21 (0.96-1.53) | 1 |
| Discharge to facility | 0.207 (0.124) | 1.23 (0.95-1.57) | 1 |

SE: standard error; CI: confidence interval
\* All risk factors are classified at the start of follow-up.

**Table 3:** Observed post-stroke cognitive impairment incidence rates by risk group in development, internal validation, and external validation cohorts.

|  | **Development** |  | **Internal Validation** |  | **External Validation** |  |
| --- | --- | --- | --- | --- | --- | --- |
| Risk category, points | Events/PY | Events/100 PY | Events/PY | Events/100 PY | Events/PY | Events/100 PY |
| Low (0-11 points) | 51/6780.5 | 0.8 | 34/3480.9 | 1.0 | 36/4403.0 | 0.8 |
| Intermediate (12-16 points) | 116/4158.8 | 2.8 | 58/2203.1 | 2.6 | 53/2244.5 | 2.4 |
| High (17-33 points) | 165/2285.1 | 7.2 | 74/1185.7 | 6.2 | 39/739.1 | 5.3 |
| Overall | 332/13,224.4 | 2.5 | 166/6869.8 | 2.4 | 128/7386.6 | 1.7 |

**Table 4:** Hazard ratios (HR) and 95% confidence intervals for incidence of post-stroke cognitive impairment in development, internal validation, and external validation cohorts.

|  | Development<br>HR (95% CI) | Internal Validation<br>HR (95% CI) | External<br>Validation<br>HR (95% CI) |
| --- | --- | --- | --- |
| High risk (12-35 points) | 9.4 (6.9-13.0) | 6.2 (4.1-9.3) | 6.1 (3.9-9.6) |
| Intermediate risk (8-11 points) | 3.7 (2.7-5.1) | 2.7 (1.8-4.1) | 2.8 (1.9-4.3) |
| Low risk (0-7 points) | - | - | - |

CI: confidence interval; HR: hazard ratio

### Internal Validation Cohort

In the internal validation cohort, there were 166 incident diagnoses of PSCI (5-year Kaplan-Meier cumulative incidence, 11.0%; 95% CI: 9.5-12.8%) and 241 death events (12.5%). The median time from the date of incident stroke to a diagnosis of PSCI was 1.9 years (IQR 0.8 – 3.3 years). The C-statistic for predicting PSCI was 0.731 (95% CI: 0.694-0.768). Like the development cohort, the observed rates of PSCI (**Table 3**) and the hazard ratios (**Table 4**) increased substantially from low to intermediate to high risk. The cumulative incidence plots also demonstrated separation by risk group (**Figure 1**).

### External Validation Cohort

In the external validation cohort, there were 128 incident diagnoses of PSCI (5-year Kaplan-Meier cumulative incidence, 7.8%; 95% CI: 6.5-9.2%) and 62 (2.8%) death events. The median time from the date of incident stroke to a diagnosis of PSCI was 1.6 years (IQR 0.8 – 2.9 years). The C-statistic was 0.724 (95% CI: 0.681-0.766). The observed rates of PSCI (**Table 3**) and HRs (**Table 4**) are lower in the external validation cohort compared to the development and internal validation cohort, but still increase substantially from low to high risk groups. Similarly, the cumulative incidence plots demonstrate separation by risk group (**Figure 1**).

## DISCUSSION

In nearly 8,000 patients from two primary care practice networks with an incident stroke, we derived and validated a model to predict the incidence of post-stroke cognitive impairment over 5 years. We observed that a model that utilizes 11 predictors that are easily ascertained from EHR data achieves good discrimination in both internal and external validation populations. A simple risk scoring scheme accurately stratifies patients into low-, intermediate-, and high-risk groups. Our findings suggest that clinical variables from electronic records ascertainable at the time of an incident stroke can be used to identify patients at increased risk of post-stroke cognitive impairment.

Prior studies have reported the results of predictive models that have moderate accuracy, but use predictors that are not readily available in clinical data.^11–14^ For example, other published models include baseline cognitive assessments (e.g. Montreal Cognitive Assessment) and/or measures of functional ability (e.g. Barthel Index, Glasgow Coma Scale) that may not be available or easily accessible for all patients.^11–14^ By using these variables that are more difficult or often unavailable in EHR data, the utility of these models is limited because of the difficulty in using them in routine clinical practice. Of the 11 features included in our final predictive model, 3 were defined from structured EHR data and 8 were defined based on ICD diagnostic billing codes Prediction models such as ours could easily be implemented and fully automated within EHRs to efficiently assess risk of PSCI at a population level.

We created a risk-scheme based on the model results which is easily calculated and classifies patients into three clinically meaningful categories. In the internal validation cohort, the “low” risk group accounted for 50% of person-years and has an observed incidence rate of PSCI of 0.98/100 person-years. The “high” risk group accounted for only 17% of person-years, but 45% of incident events and an observed incidence rate of PSCI of 6.24/100 person-years. This risk scheme may be clinically applicable in efficiently identifying patients at highest risk who would be more likely to benefit from neuropsychiatric and social interventions, as well as interventions targeted at modifiable risk factors to limit PSCI.

Many published models predicting PSCI have short-term follow-up of only 3-6 months following the stroke event and may be mostly capturing PSCI that is directly related to the stroke itself. Although the rate of PSCI is highest shortly after the stroke, the cognitive impairment that occurs in the first few months of the stroke may be reversible.^28^ Many patients experience delayed-onset PSCI which occurs >6 months after the stroke.^29^ Our model predicts PSCI within 5 years when beginning follow-up 3 months following the stroke to remove PSCI directly related to the stroke itself, which provides an opportunity for targeted secondary prevention interventions in high-risk patients. We also focused on patients experiencing their first stroke documented in the EHR. A major risk factor for PSCI is a recurrent stroke so targeting those at increased risk for PSCI may be a strategy to reduce both secondary stroke events and PSCI.^3^

Our model performed favorably in an external validation population which was younger and more ethnically diverse, and used more recent electronic health record data than our model derivation population. Although the external validation population was a lower risk group and may represent less severe strokes, the model still had good discrimination with a c-statistic of 0.724. Additionally, the 3-category risk scheme was able to separate “low” risk patients (incidence rate of 0.82/100 person-years) from “high” risk patients (incidence rate of 5.28/100 person-years).

This study has several potential limitations. This model was developed using patients from one primary care practice network affiliated with an academic medical center that were largely of European ancestry, so generalizability may be limited. However, the model still performed well in an external population that was younger and more ethnically diverse. Ascertainment of clinical features and incidence of PSCI was based on retrospective assessment of EHR documentation, which may be associated with misclassification. Data on clinical features and ascertainment of both incident and prevalent strokes and cognitive impairment is limited to what is available within the Mass General Brigham EHR. We do not have the ability to fully ascertain information on clinical features or incident diagnoses of stroke/cognitive impairment for patients seen outside of our network. We were not able to evaluate the severity of stroke events since many occurred outside our network. It is also possible that prior strokes occurring outside of our network or prior to enrollment may not have been documented, especially if mild or with resolution of symptoms.

In conclusion, we developed and validated a prediction model of five-year risk of PSCI in a primary care population using routinely collected electronic health record data. Model output categorized patients into three risk groups that could be used to identify individuals at increased risk for post-stroke cognitive impairment for preventive efforts.

## Supplementary Material

Table S1-S4

## Data Availability

Mass General Brigham data contains protected health information and cannot be shared publicly. The data processing scripts used to perform analyses will be made available to interested researchers upon reasonable request to the corresponding author.

