## Supplemental Table 1-4 for "Predicting post-stroke cognitive impairment using electronic health record data"

**Table S1: ICD-9 and ICD-10 codes used to identify diagnoses of stroke**

| Ischemic Stroke | | |
|---|---|---|
| Code | Code Type | Description |
| I63 | ICD10 | Cerebral infarction |
| I63.00 | ICD10 | Cerebral infarction due to thrombosis of unspecified precerebral artery |
| I63.011 | ICD10 | Cerebral infarction due to thrombosis of right vertebral artery |
| I63.012 | ICD10 | Cerebral infarction due to thrombosis of left vertebral artery |
| I63.013 | ICD10 | Cerebral infarction due to thrombosis of bilateral vertebral arteries |
| I63.019 | ICD10 | Cerebral infarction due to thrombosis of unspecified vertebral artery |
| I63.02 | ICD10 | Cerebral infarction due to thrombosis of basilar artery |
| I63.031 | ICD10 | Cerebral infarction due to thrombosis of right carotid artery |
| I63.032 | ICD10 | Cerebral infarction due to thrombosis of left carotid artery |
| I63.033 | ICD10 | Cerebral infarction due to thrombosis of bilateral carotid arteries |
| I63.039 | ICD10 | Cerebral infarction due to thrombosis of unspecified carotid artery |
| I63.09 | ICD10 | Cerebral infarction due to thrombosis of other precerebral artery |
| I63.10 | ICD10 | Cerebral infarction due to embolism of unspecified precerebral artery |
| I63.111 | ICD10 | Cerebral infarction due to embolism of right vertebral artery |
| I63.112 | ICD10 | Cerebral infarction due to embolism of left vertebral artery |
| I63.113 | ICD10 | Cerebral infarction due to embolism of bilateral vertebral arteries |
| I63.119 | ICD10 | Cerebral infarction due to embolism of unspecified vertebral artery |
| I63.12 | ICD10 | Cerebral infarction due to embolism of basilar artery |
| I63.131 | ICD10 | Cerebral infarction due to embolism of right carotid artery |
| I63.132 | ICD10 | Cerebral infarction due to embolism of left carotid artery |
| I63.133 | ICD10 | Cerebral infarction due to embolism of bilateral carotid arteries |
| I63.139 | ICD10 | Cerebral infarction due to embolism of unspecified carotid artery |
| I63.19 | ICD10 | Cerebral infarction due to embolism of other precerebral artery |
| I63.20 | ICD10 | Cerebral infarction due to unspecified occlusion or stenosis of unspecified precerebral arteries |
| I63.211 | ICD10 | Cerebral infarction due to unspecified occlusion or stenosis of right vertebral artery |
| I63.212 | ICD10 | Cerebral infarction due to unspecified occlusion or stenosis of left vertebral arteries |
| I63.213 | ICD10 | Cerebral infarction due to unspecified occlusion or stenosis of bilateral vertebral arteries |
| I63.219 | ICD10 | Cerebral infarction due to unspecified occlusion or stenosis of unspecified vertebral arteries |
| I63.22 | ICD10 | Cerebral infarction due to unspecified occlusion or stenosis of basilar arteries |
| I63.231 | ICD10 | Cerebral infarction due to unspecified occlusion or stenosis of right carotid arteries |
| I63.232 | ICD10 | Cerebral infarction due to unspecified occlusion or stenosis of left carotid arteries |

| | | |
|---|---|---|
| I63.233 | ICD10 | Cerebral infarction due to unspecified occlusion or stenosis of bilateral carotid arteries |
| I63.239 | ICD10 | Cerebral infarction due to unspecified occlusion or stenosis of unspecified carotid arteries |
| I63.29 | ICD10 | Cerebral infarction due to unspecified occlusion or stenosis of other precerebral arteries |
| I63.30 | ICD10 | Cerebral infarction due to thrombosis of unspecified cerebral artery |
| I63.311 | ICD10 | Cerebral infarction due to thrombosis of right middle cerebral artery |
| I63.312 | ICD10 | Cerebral infarction due to thrombosis of left middle cerebral artery |
| I63.313 | ICD10 | Cerebral infarction due to thrombosis of bilateral middle cerebral arteries |
| I63.319 | ICD10 | Cerebral infarction due to thrombosis of unspecified middle cerebral artery |
| I63.321 | ICD10 | Cerebral infarction due to thrombosis of right anterior cerebral artery |
| I63.322 | ICD10 | Cerebral infarction due to thrombosis of left anterior cerebral artery |
| I63.323 | ICD10 | Cerebral infarction due to thrombosis of bilateral anterior cerebral arteries |
| I63.329 | ICD10 | Cerebral infarction due to thrombosis of unspecified anterior cerebral artery |
| I63.331 | ICD10 | Cerebral infarction due to thrombosis of right posterior cerebral artery |
| I63.332 | ICD10 | Cerebral infarction due to thrombosis of left posterior cerebral artery |
| I63.333 | ICD10 | Cerebral infarction due to thrombosis of bilateral posterior cerebral arteries |
| I63.339 | ICD10 | Cerebral infarction due to thrombosis of unspecified posterior cerebral artery |
| I63.341 | ICD10 | Cerebral infarction due to thrombosis of right cerebellar artery |
| I63.342 | ICD10 | Cerebral infarction due to thrombosis of left cerebellar artery |
| I63.343 | ICD10 | Cerebral infarction to thrombosis of bilateral cerebellar arteries |
| I63.349 | ICD10 | Cerebral infarction due to thrombosis of unspecified cerebellar artery |
| I63.39 | ICD10 | Cerebral infarction due to thrombosis of other cerebral artery |
| I63.40 | ICD10 | Cerebral infarction due to embolism of unspecified cerebral artery |
| I63.411 | ICD10 | Cerebral infarction due to embolism of right middle cerebral artery |
| I63.412 | ICD10 | Cerebral infarction due to embolism of left middle cerebral artery |
| I63.413 | ICD10 | Cerebral infarction due to embolism of bilateral middle cerebral arteries |
| I63.419 | ICD10 | Cerebral infarction due to embolism of unspecified middle cerebral artery |
| I63.421 | ICD10 | Cerebral infarction due to embolism of right anterior cerebral artery |
| I63.422 | ICD10 | Cerebral infarction due to embolism of left anterior cerebral artery |
| I63.423 | ICD10 | Cerebral infarction due to embolism of bilateral anterior cerebral arteries |
| I63.429 | ICD10 | Cerebral infarction due to embolism of unspecified anterior cerebral artery |
| I63.431 | ICD10 | Cerebral infarction due to embolism of right posterior cerebral artery |
| I63.432 | ICD10 | Cerebral infarction due to embolism of left posterior cerebral artery |
| I63.433 | ICD10 | Cerebral infarction due to embolism of bilateral posterior cerebral arteries |
| I63.439 | ICD10 | Cerebral infarction due to embolism of unspecified posterior cerebral artery |
| I63.441 | ICD10 | Cerebral infarction due to embolism of right cerebellar artery |

| | | |
|---|---|---|
| I63.442 | ICD10 | Cerebral infarction due to embolism of left cerebellar artery |
| I63.443 | ICD10 | Cerebral infarction due to embolism of bilateral cerebellar arteries |
| I63.449 | ICD10 | Cerebral infarction due to embolism of unspecified cerebellar artery |
| I63.49 | ICD10 | Cerebral infarction due to embolism of other cerebral artery |
| I63.50 | ICD10 | Cerebral infarction due to unspecified occlusion or stenosis of unspecified cerebral artery |
| I63.511 | ICD10 | Cerebral infarction due to unspecified occlusion or stenosis of right middle cerebral artery |
| I63.512 | ICD10 | Cerebral infarction due to unspecified occlusion or stenosis of left middle cerebral artery |
| I63.513 | ICD10 | Cerebral infarction due to unspecified occlusion or stenosis of bilateral middle cerebral arteries |
| I63.519 | ICD10 | Cerebral infarction due to unspecified occlusion or stenosis of unspecified middle cerebral artery |
| I63.521 | ICD10 | Cerebral infarction due to unspecified occlusion or stenosis of right anterior cerebral artery |
| I63.522 | ICD10 | Cerebral infarction due to unspecified occlusion or stenosis of left anterior cerebral artery |
| I63.523 | ICD10 | Cerebral infarction due to unspecified occlusion or stenosis of bilateral anterior cerebral arteries |
| I63.529 | ICD10 | Cerebral infarction due to unspecified occlusion or stenosis of unspecified anterior cerebral artery |
| I63.531 | ICD10 | Cerebral infarction due to unspecified occlusion or stenosis of right posterior cerebral artery |
| I63.532 | ICD10 | Cerebral infarction due to unspecified occlusion or stenosis of left posterior cerebral artery |
| I63.533 | ICD10 | Cerebral infarction due to unspecified occlusion or stenosis of bilateral posterior cerebral arteries |
| I63.539 | ICD10 | Cerebral infarction due to unspecified occlusion or stenosis of unspecified posterior cerebral artery |
| I63.541 | ICD10 | Cerebral infarction due to unspecified occlusion or stenosis of right cerebellar artery |
| I63.542 | ICD10 | Cerebral infarction due to unspecified occlusion or stenosis of left cerebellar artery |
| I63.543 | ICD10 | Cerebral infarction due to unspecified occlusion or stenosis of bilateral cerebellar arteries |
| I63.549 | ICD10 | Cerebral infarction due to unspecified occlusion or stenosis of unspecified cerebellar artery |
| I63.59 | ICD10 | Cerebral infarction due to unspecified occlusion or stenosis of other cerebral artery |
| I63.6 | ICD10 | Cerebral infarction due to cerebral venous thrombosis, nonpyogenic |
| I63.8 | ICD10 | Other cerebral infarction |
| I63.81 | ICD10 | Other cerebral infarction due to occlusion or stenosis of small artery |
| I63.89 | ICD10 | Other cerebral infarction |
| I63.9 | ICD10 | Cerebral infarction, unspecified |
| 433.01 | ICD9 | Occlusion and stenosis of basilar artery with cerebral infarction |
| 433.11 | ICD9 | Occlusion and stenosis of carotid artery with cerebral infarction |

| | | |
|---|---|---|
| 433.21 | ICD9 | Occlusion and stenosis of vertebral artery with cerebral infarction |
| 433.31 | ICD9 | Occlusion and stenosis of multiple and bilateral precerebral arteries with cerebral infarction |
| 433.81 | ICD9 | Occlusion and stenosis of other specified precerebral artery with cerebral infarction |
| 433.91 | ICD9 | Occlusion and stenosis of unspecified precerebral artery with cerebral infarction |
| 434.01 | ICD9 | Cerebral thrombosis with cerebral infarction |
| 434.11 | ICD9 | Cerebral embolism with cerebral infarction |
| 434.91 | ICD9 | Cerebral artery occlusion, unspecified with cerebral infarction |
| Hemorrhagic Stroke – Used for exclusions only | | |
| I60 | ICD10 | Nontramatic subarchnoid haemorrhage |
| I60.00 | ICD10 | Nontraumatic subarachnoid hemorrhage from unspecified carotid siphon and bifurcation |
| I60.01 | ICD10 | Nontraumatic subarachnoid hemorrhage from right carotid siphon and bifurcation |
| I60.02 | ICD10 | Nontraumatic subarachnoid hemorrhage from left carotid siphon and bifurcation |
| I60.10 | ICD10 | Nontraumatic subarachnoid hemorrhage from unspecified middle cerebral artery |
| I60.11 | ICD10 | Nontraumatic subarachnoid hemorrhage from right middle cerebral artery |
| I60.12 | ICD10 | Nontraumatic subarachnoid hemorrhage from left middle cerebral artery |
| I60.2 | ICD10 | Nontraumatic subarachnoid hemorrhage from left carotid siphon and bifurcation |
| I60.20 | ICD10 | Nontraumatic subarachnoid hemorrhage from unspecified anterior communicating artery |
| I60.21 | ICD10 | Nontraumatic subarachnoid hemorrhage from right anterior communicating artery |
| I60.22 | ICD10 | Nontraumatic subarachnoid hemorrhage from left anterior communicating artery |
| I60.30 | ICD10 | Nontraumatic subarachnoid hemorrhage from unspecified posterior communicating artery |
| I60.31 | ICD10 | Nontraumatic subarachnoid hemorrhage from right posterior communicating artery |
| I60.32 | ICD10 | Nontraumatic subarachnoid hemorrhage from left posterior communicating artery |
| I60.4 | ICD10 | Nontraumatic subarachnoid hemorrhage from basilar artery |
| I60.50 | ICD10 | Nontraumatic subarachnoid hemorrhage from unspecified vertebral artery |
| I60.51 | ICD10 | Nontraumatic subarachnoid hemorrhage from right vertebral artery |

| | | |
|---|---|---|
| I60.52 | ICD10 | Nontraumatic subarachnoid hemorrhage from left vertebral artery |
| I60.6 | ICD10 | Nontraumatic subarachnoid hemorrhage from other intracranial arteries |
| I60.7 | ICD10 | Nontraumatic subarachnoid hemorrhage from unspecified intracranial artery |
| I60.8 | ICD10 | Other nontraumatic subarachnoid hemorrhage |
| I60.9 | ICD10 | Nontraumatic subarachnoid hemorrhage, unspecified |
| I61 | ICD10 | Nontraumatic intracerebral haemorrage |
| I61.0 | ICD10 | Nontraumatic intracerebral hemorrhage in hemisphere, subcortical |
| I61.1 | ICD10 | Nontraumatic intracerebral hemorrhage in hemisphere, cortical |
| I61.2 | ICD10 | Nontraumatic intracerebral hemorrhage in hemisphere, unspecified |
| I61.3 | ICD10 | Nontraumatic intracerebral hemorrhage in brain stem |
| I61.4 | ICD10 | Nontraumatic intracerebral hemorrhage in cerebellum |
| I61.5 | ICD10 | Nontraumatic intracerebral hemorrhage, intraventricular |
| I61.6 | ICD10 | Nontraumatic intracerebral hemorrhage, multiple localized |
| I61.8 | ICD10 | Other nontraumatic intracerebral hemorrhage |
| I61.9 | ICD10 | Nontraumatic intracerebral hemorrhage, unspecified |
| I62 | ICD10 | Other and unspecified nontraumatic intracranial hemorrhage |
| I62.0 | ICD10 | Nontraumatic subdural hemorrhage |
| I62.00 | ICD10 | Nontraumatic subdural hemorrhage unspecified |
| I62.01 | ICD10 | Nontraumatic acute subdural hemorrhage |
| I62.02 | ICD10 | Nontraumatic subacute subdural hemorrhage |
| I62.03 | ICD10 | Nontraumatic chronic subdural hemorrhage |
| I62.1 | ICD10 | Nontraumatic extradural hemorrhage |
| I62.9 | ICD10 | Nontraumatic intracranial hemorrhage, unspecified |
| 430 | ICD9 | Subarachnoid hemorrhage |
| 431 | ICD9 | Intracerebral hemorrhage |
| 432 | ICD9 | Other and unspecified intracranial hemorrhage |
| 432.9 | ICD9 | Unspecified intracranial hemorrhage |

**Table S2. List of clinical features considered in model to predict post-stroke cognitive impairment**

| Variable | Data Source |
|---|---|
| **Patient characteristics** | |
| Age | Demographics |
| Sex | Demographics |
| Insurance | Demographics |
| Marital status | Demographics |
| Education level | Demographics |
| Income < poverty level | Geocoding |
| Income < $30,000 | Geocoding |
| Income < $50,000 | Geocoding |
| Current smoker | EHR structured field |
| Alcohol abuse | ICD |
| Physical inactivity | Unstructured Notes |
| Discharge to facility | Discharge Summary |
| **Comorbidities** | |
| Anxiety | ICD |
| Chronic kidney disease – severe | Validated Algorithm |
| Coronary artery disease | Validated Algorithm |
| Delirium | ICD |
| Depression | ICD |
| Hyperlipidemia | ICD |
| Obesity | Validated Algorithm |
| **Electronic frailty index (eFI) factors*** | |
| Activity limitation | |
| Anemia | ICD |
| Arthritis | ICD |
| Atrial fibrillation | Validated Algorithm |
| Cerebrovascular disease | Validated Algorithm |
| Chronic kidney disease | Validated Algorithm |
| Congestive heart failure | Validated Algorithm |
| Diabetes | Validated Algorithm |
| Dizziness | ICD |
| Dyspnea | ICD |
| Falls | ICD |
| Foot problems | ICD |
| Fragility fracture | ICD |
| Hearing impairment | ICD |
| Heart valve disease | ICD |
| Housebound | ICD |
| Hypertension | Validated Algorithm |
| Hypotension/syncope | ICD |
| Ischemic heart disease | ICD |

| | |
|---|---|
| Mobility problems / functional limitations | ICD |
| Osteoporosis | ICD |
| Parkinsonism | ICD |
| Peptic ulcer | ICD |
| Peripheral vascular disease | Validated Algorithm |
| Polypharmacy | Medication |
| Requirement for care | ICD |
| Respiratory disease | ICD |
| Skin ulcer | ICD |
| Sleep disturbances | ICD + Medication |
| Social vulnerability | ICD |
| Thyroid disease | ICD |
| Urinary incontinence | ICD |
| Urinary system disease | ICD |
| Visual impairment | ICD |
| Weight loss and anorexia | ICD |
| **Laboratory values + vital signs** | |
| Estimated glomerular filtration rate (eGFR) | Laboratory |
| Hemoglobin A1c (HbA1c) | Laboratory |
| HDL cholesterol | Laboratory |
| LDL cholesterol | Laboratory |
| Total cholesterol | Laboratory |
| Triglycerides | Laboratory |
| Systolic blood pressure | Vital signs |
| **Medications** | |
| Anti-hypertensives | Medication |
| Anticoagulants | Medication |
| Antiplatelets | Medication |
| Aspirin | Medication |
| Statins | Medication |
| **Neurologic** | |
| Ventricular enlargement | MRI Report |
| Traumatic brain injury | ICD |
| White matter disease | MRI Report |
| White matter infarct | MRI Report |

* eFI is an electronic frailty index containing 36 deficits. We considered each deficit individually, instead of as a composite score. “Memory and cognitive problems” was not considered as a predictor, so 35 of the 36 deficits were included.

**Table S3: Regular expression coding of brain MRI reports**

| Variable | Regular Expression Terms |
|---|---|
| White matter disease | Notes of "T2/Flair hyperintensity", "T2/Flair hyperintensities", "T2 hyperintensity", "T2 hyperintensities" followed by a mention of any of the following words within the next 100 characters: “periventricular”, “deep white matter” “subcortical white matter”.<br>Negation: “no evidence of significant white matter disease“, “no evidence of white matter disease”, “no evidence of significant periventricular deep white matter disease“, “no evidence of periventricular deep white matter disease“, “no evidence of periventricular white matter disease“, “minimal white matter disease”. |
| White matter infarcts | Notes of “DEEP WHITE”, “WHITE MATTER LESION”,“WHITE MATTER INFARCTION”, “WHITE MATTER INFARCT”, or “WHITE MATTER DISEASE” followed by the word “INFARCT”<br>Negation: 'no evidence of acute infarction', 'no acute infarct', 'no evidence of intracranial mass or acute infarction', 'no evidence of territorial infarct', 'there is no restricted diffusion to suggest acute infarction', 'no infarction is identified', 'no infarct', 'no restricted diffusion to suggest acute infarction', 'no evidence of areas of restricted diffusion to suggest an acute infarct'. Also, any occurrence of the words “no evidence of”, “without evidence of”, “minimal” within 200 characters of the main White Matter Infarct flag is also considered a negation. |
| Ventricular enlargement | “ventricular enlargement”, “the size of the ventricles is enlarged”, “enlarged ventricles”, “enlargement of ventricles”, “enlargement of the ventricles”.<br>Negation: any occurrence of the following expressions 'no evidence of ventricular enlargement', 'no ventricular enlargement',<br>'no evidence of ventricular enlargement'<br>'no evidence of enlargement of the ventricles'<br>'there is no enlargement of the ventricles'<br>'without evidence of ventricular enlargement'<br>'without evidence of enlargement of the ventricles' |

**Table S4: ICD-9 and ICD-10 codes used to identify diagnoses of dementia and cognitive impairment**

| Code type | Code | Description | Notes |
|---|---|---|---|
| ICD-10 | F01 | Vascular dementia | |
| ICD-10 | F01.5 | Vascular dementia | |
| ICD-10 | F01.50 | Vascular dementia without behavioral disturbance | |
| ICD-10 | F01.51 | Vascular dementia with behavioral disturbance | |
| ICD-10 | F02 | Dementia in other diseases classified elsewhere | |
| ICD-10 | F02.8 | Dementia in other diseases classified elsewhere | |
| ICD-10 | F02.80 | Dementia in other diseases classified elsewhere without behavioral disturbance | |
| ICD-10 | F02.81 | Dementia in other diseases classified elsewhere with behavioral disturbance | |
| ICD-10 | F03 | Unspecified dementia | |
| ICD-10 | F03.9 | Unspecified dementia | |
| ICD-10 | F03.90 | Unspecified dementia without behavioral disturbance | |
| ICD-10 | F03.91 | Unspecified dementia with behavioral disturbance | |
| ICD-10 | G23.1 | Progressive supranuclear ophthalmoplegia (Steele-Richardson-Olszewski) | |
| ICD-10 | G30.0 | Alzheimer's disease with early onset | |
| ICD-10 | G30.1 | Alzheimer's disease with late onset | |
| ICD-10 | G30.8 | Other Alzheimer's disease | |
| ICD-10 | G30.9 | Alzheimer's disease, unspecified | |
| ICD-10 | G31.0 | Frontotemporal dementia | |
| ICD-10 | G31.01 | Pick's Disease | |
| ICD-10 | G31.09 | Other frontotemporal dementia | |
| ICD-10 | G31.1 | Senile degeneration of brain, not elsewhere classified | |
| ICD-10 | G31.83 | Dementia with Lewy bodies | |
| ICD-10 | G31.84 | Mild cognitive impairment, so stated | |
| ICD-10 | G21.4 | Vascular parkinsonism | |
| ICD-10 | I69.01 | Cognitive deficits following nontraumatic subarachnoid hemorrhage | |
| ICD-10 | I69.11 | Cognitive deficits following nontraumatic intracerebral hemorrhage | |
| ICD-10 | I69.21 | Cognitive deficits following other nontraumatic intracranial hemorrhage | |
| ICD-10 | I69.31 | Cognitive deficits following cerebral infarction | |

| | | | |
|---|---|---|---|
| ICD-10 | I69.81 | Cognitive deficits following other cerebrovascular disease | |
| ICD-10 | I69.91 | Cognitive deficits following unspecified cerebrovascular disease | |
| ICD-10 | F10.27 | Alcohol dependence with other alcohol-induced disorders with alcohol-induced persisting dementia | Used to identify prevalent disease only |
| ICD-10 | F10.97 | Alcohol use, unspecified with alcohol-induced psychotic disorder with alcohol-induced persisting dementia | Used to identify prevalent disease only |
| ICD-9 | 290.0 | Senile dementia, uncomplicated | |
| ICD-9 | 290.1 | Presenile dementia, uncomplicated | |
| ICD-9 | 290.11 | Presenile dementia with delirium | |
| ICD-9 | 290.12 | Presenile dementia with delusional features | |
| ICD-9 | 290.13 | Presenile dementia with depressive features | |
| ICD-9 | 290.2 | Senile dementia with delusional features | |
| ICD-9 | 290.21 | Senile dementia with depressive features | |
| ICD-9 | 290.3 | Senile dementia with delirium | |
| ICD-9 | 290.4 | Vascular dementia, uncomplicated | |
| ICD-9 | 290.41 | Vascular dementia, with delirium | |
| ICD-9 | 290.42 | Vascular dementia, with delusions | |
| ICD-9 | 290.43 | Vascular dementia, with depressed mood | |
| ICD-9 | 290.8 | Other specified senile psychotic conditions | |
| ICD-9 | 290.9 | Unspecified senile psychotic condition | |
| ICD-9 | 294.1 | Dementia in conditions classified elsewhere | |
| ICD-9 | 294.10 | Dementia in conditions classified elsewhere without behavioral disturbance | |
| ICD-9 | 294.11 | Dementia in conditions classified elsewhere with behavioral disturbance | |
| ICD-9 | 294.20 | Dementia, unspecified, without behavioral disturbance | |
| ICD-9 | 294.21 | Dementia, unspecified, with behavioral disturbance | |
| ICD-9 | 331.0 | Alzheimer's disease | |
| ICD-9 | 331.1 | Frontotemporal dementia | |
| ICD-9 | 331.11 | Pick's disease | |
| ICD-9 | 331.19 | Other frontotemporal dementia | |
| ICD-9 | 331.2 | Senile degeneration of brain | |
| ICD-9 | 331.82 | Dementia with Lewy bodies | |
| ICD-9 | 331.83 | Mild cognitive impairment | |
| ICD-9 | 331.9 | Cerebral degeneration unspecified | |

| | | | |
|---|---|---|---|
| ICD-9 | 438.0 | Late effects of cerebrovascular disease, cognitive deficits | |
| ICD-9 | 291.2 | Alcohol-induced persisting dementia | Used to identify prevalent disease only |
